# Family Planning Performance in Selected States and Union Territories of India: Evidence from NFHS 2019-20

**DOI:** 10.1101/2020.12.17.20248407

**Authors:** Aalok Ranjan Chaurasia

**Affiliations:** MLC Foundation and ‘Shyam’ Institute Bhopal, MP, India

**Keywords:** India, States, Union Territories, Family Planning, Performance, Composite Index

## Abstract

This paper uses a new composite index to measure family planning performance in 22 states/Union Territories of India for which latest data on family planning use are available from NFHS 2019-2020. The analysis reveals that family planning performance in meeting the family planning needs of the people remains poor and there are significant inter-state/Union Territory variation in the performance. The paper calls for reinvigorating family planning efforts by treating family planning as a development strategy.

## Introduction

Organised family planning efforts in India date back to 1952 when world’s first official Family Planning Programme was launched. In its initial phase, the programme was directed towards improving health and welfare of the family, especially children and women (Chaurasia and Singh, 2014). Subsequently, it focused on birth limitation to curb population growth and a target-based approach was adopted for programme implementation. Programme performance, in this approach, was measured in terms of equivalent sterilisations and couples effectively protected (Chaurasia, 1985; Government of India, 1990). In 1969, the first nationally representative family planning survey was conducted which revealed that only 14 per cent currently married women aged 15-44 years were using a family planning method while less than 10 per cent were using a modern method (Operations Research Group, 1970). The second all India survey, carried out in 1980, revealed that family planning use among currently married women aged 15-44 years was around 35 per cent with 28 per cent women using a modern method (Khan and Prasad, 1980). The first National Family Health Survey (NFHS) 1992-93 revealed a contraceptive prevalence rate (CPR) – proportion of currently married women aged 15-49 using a family planning method - of 40 per cent while the prevalence of modern family planning methods (mCPR) was 36 per cent (International Institute for Population Sciences, 1995). In 1996, Government of India abolished the target-based approach of implementation and CPR and mCPR became the basis for measuring family planning performance. NFHS 1998-99 revealed that CPR increased to 45 per cent (International Institute for Population Sciences and ORC Macro, 2000) while NFHS 2005-06 estimated a CPR of 55 per cent (International Institute for Population Sciences and Macro International, 2007). However, NFHS 2015-16, reported a decrease in CPR to 53 per cent while mCPR stagnated at around 48 per cent with substantial within country variations country (International Institute for Population Sciences and ICF, 2017).

The popularity of CPR, as an indicator of family planning performance, rests with its strong inverse relationship with the total fertility rate (TFR) on the basis of cross-country data (Bongaarts, 1978; Bongaarts and Potter, 1983; Ross and Mauldin, 1996; Jain, 1997; Tsui, 2001; Stover, 1998; United Nations, 2020). There are, however, studies that show inconsistency between CPR and TFR in many countries (United Nations, 2020). Srinivasan (1988) has argued that as one goes down the level of aggregation, variation in family planning use explains less and less of the variation in TFR. Chaurasia (2004) has found that the variation in family planning use explains only around 20 per cent of the variation in the total marital fertility rate below the district level in Madhya Pradesh. Chaurasia (2000) has also observed that the number of children ever born in Madhya Pradesh was positively related to family planning use at the individual level.

CPR, however, is not an appropriate indicator to measure family planning performance for many reasons. CPR is essentially a ratio, not the rate or the incidence of family planning practice. It is a crude indicator that does not take into consideration the variation in the use of different family planning methods. The upper limit of CPR is also difficult to establish as a substantial proportion of currently married reproductive age women do not practice family planning because they either want child or they are pregnant or the woman or her partner is sterile and this proportion varies widely across different population groups. For all practical purposes, CPR can never be 100 per cent.

It is well-known that family planning needs of a couple vary by different phases of the family building process and are conditioned by such factors as personal circumstances, individual knowledge and changing childbearing preferences. Availability and accessibility of different family planning methods and their effectiveness also affect use of different family planning methods. Family planning allows couples to anticipate and attain their desired number of children and the spacing and timing of their births which has a direct impact on health and well-being of women and the outcome of pregnancy. It has, therefore, been emphasised that family planning performance measurement must also take into account the range and types of family planning methods being used (United Nations, 2019). This means that family planning performance should be measured on a two-dimensional space – the dimension of the met demand for family planning and the dimension of the structure or composition of the met demand. The CPR neither captures the met demand for family planning nor the composition of the met demand.

Recently, Chaurasia (2020) has developed a composite family planning performance index that takes into account both the met demand for family planning and the structure of the met demand. The index is based on the underlying principles of informed choice and provision of a range of family planning methods to potential family planning users so as to meet the diverse family planning needs of the people to plan their family. The index is similar to the multi-dimensional poverty index that has been proposed by Anand and Sen (1997) to measure poverty.

The aim of his paper is to apply composite family planning performance index proposed by Chaurasia (2020) to measure family planning performance in 22 states/Union Territories of the country for which the latest data on the use of different family planning methods and the unmet need of family planning for spacing and for stopping births are available through the latest NFHS carried out during 2019-20. The paper also analyses the variation in the improvement in family planning performance across different states and Union Territories during the period 2015-16 and 2019-20. The analysis presents a different picture of family planning performance than that revealed through CPR.

Family Planning Performance Index

The family planning performance index, *p*, is defined as (Chaurasia, 2020):

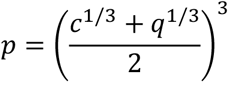

Where the index *c* measures the met demand for family planning and the index *q* measures the structure of the met demand. The index *c* is defined as

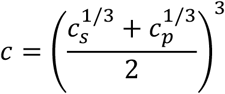

Where the index *c*_*s*_ measures the met demand for modern spacing methods and the index *c*_*p*_ measures the met demand for permanent methods. The index *c*_*s*_ is defined as

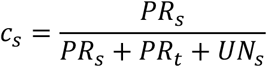

where *PR*_*s*_ is the prevalence of modern spacing methods, *PR*_*t*_ is the prevalence of traditional methods and *UN*_*s*_ is the unmet need for spacing methods. It is assumed that the use of traditional methods actually reflects the unmet demand for spacing methods. On the other hand, the index *c*_*p*_ is defined as

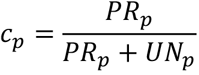

where *PR*_*p*_ is the prevalence of permanent methods and *UN*_*p*_ is the unmet need for permanent methods. Both *c*_*s*_ and *c*_*p*_ vary between 0 and 1 and the higher the index the higher the met demand.

On the other hand, the index *q* is defined as

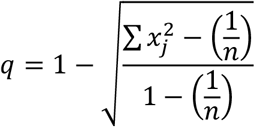

where *x*_*j*_ is the proportionate prevalence of the contraceptive method *j* and *n* is the number of contraceptive methods available. The index *q* is essentially a measure of the skewness in contraceptive method mix. The lower the index the higher the method skew and vice versa. A skewed method mix is a reflection that family planning services delivery system is not able to meet the diverse family planning needs of the people. When the entire family planning use is confined to only one method *q*=0. When family planning use is evenly distributed across different family planning methods available, *q*=1. The rationale behind the construction of the index *p* is given elsewhere (Chaurasia, 2020). The index varies between 0 and 1 and the higher the index the better the performance. Family planning performance may be rated as very poor if *p*<0.300; poor if 0.300≤*p*<0.550; average if 0.550≤*p*<0.750; good if 0.750≤*p*<0.900; and very good if *p*≥0.900.

### Data Source

The analysis is based on estimates of method-specific prevalence rates for six modern family planning methods – female sterilisation, male sterilisation, IUD including post-partum IUD, pill, condom and injectable – and the unmet need of spacing and stopping births for 22 states/Union Territories of the country recently released by the Government of India on the basis of the latest NFHS 2019-20 (Government of India, 2020). Estimates of the prevalence of all methods of family planning combined are also available for these states from NFHS 2019-20 which allows estimation of the prevalence of traditional methods of family planning. Data related to the prevalence of different methods of family planning and the unmet need for spacing and stopping births for other states and Union Territories as well as for the country as a whole from NFHS 2019-20 have not yet been released by the Government of India. NFHS is the only source of information related to the extent of the use of different family planning methods in the country and in its constituent states, Union Territories and districts. Estimates of method-specific prevalence rates available through NFHS cover both services provided under the official family planning activities and non-programme family planning services. Family planning practice in India, however, is largely a prerogative of official family planning efforts so that analysis of family planning performance on the basis of the data available through NFHS largely reflects the performance of official family planning efforts.

### Family Planning Performance

The data available through NFHS 2019-20 suggest that family planning performance among the 22 states/Union Territories is relatively the best in the Union Territory of Laddakh but relatively the poorest in Andhra Pradesh. Laddakh is the only state/Union Territory of the country where family planning performance can be rated as very good. On the other hand, in 7 states/Union Territories, family planning performance can be rated as average and in 11 states/Union Territories, the performance can be rated as poor. This leaves three states – Kerala, Telangana and Andhra Pradesh – where family planning performance can be rated as very poor.

The index *p* is a composite of the indexes *c* and *q*. The index *c* is estimated to be the highest in Karnataka closely followed by Laddakh. There are 6 states/Union Territories where more than 75 per cent of the total demand for modern family planning methods is met whereas the met demand for modern family planning methods is less than 30 per cent in Manipur. In 7 states/Union Territories, only 60-70 per cent of the total demand for modern family planning methods is met whereas in 5 states/Union Territories, the met demand for modern family planning methods ranges between 40-50 per cent. There is no state/Union Territory where the met demand for modern family planning methods is at least 90 per cent.

The met demand for modern family planning methods is determined by the met demand for modern spacing methods and the met demand for permanent methods which are captured through indexes *c*_*s*_ and *c*_*p*_. The met demand for modern spacing methods is less than 30 per cent in five states/Union Territories whereas there is no state/Union Territory where the met demand for permanent methods is less than 30 per cent. In 15 states/Union Territories, the met demand for permanent methods is more than 80 per cent and, in three states, it is more than 90 per cent. By contrast, the met demand for modern spacing methods is more than 80 per cent in the Union Territory of Laddakh only. There is no state/Union Territory where the met demand for modern spacing methods is at least 90 per cent.

On the other hand, the index *q* is the highest in Laddakh but the lowest in Andhra Pradesh. In Andhra Pradesh, female sterilization, alone, accounts for more than 98 per cent of the total family planning use so that the method mix is extremely skewed and the index *q* is very close to 0 which reflects very poor capacity of the family planning services delivery system in the state to meet the diverse family planning needs of the people. Other states where the index *q* is very low are Karnataka, Kerala and Telangana. By contrast, the index *q* is high in Sikkim, Manipur and Jammu and Kashmir. In these states and Union Territories, the method mix is less skewed which implies that the family planning services delivery system is able to meet most of the diverse family planning needs of the people.

The classification of states/Union Territories on the two-dimensional space suggests that in majority of the states, the met demand for modern family planning methods may be rated as average while the composition of the met demand may be rated as poor (Table 3). There is no state/Union Territories where both the met demand for modern family planning methods and the composition of the met demand may be rated as very good. Similarly, there is no state/Union Territory where both met demand and composition of the met demand may be rate as very poor. In 6 states/Union Territories, the met demand for modern family planning methods may be rated as good but the composition of the met demand varies from very poor to average. Similarly, there are 4 states/Union Territories where the composition of the met demand may be rated as average but the met demand varied from very poor to good. There are only two Union Territories where the met demand for modern family planning methods is rated as good and the composition of the met demand is rated as average. In these Union Territories, the skewness in the method mix is the lowest and the family planning performance is relatively the best. On the other hand, there are states where the met demand for modern family planning methods is poor and the composition of the met demand is very poor. In these states, the method mix is extremely skewed which implies that the family planning needs of a substantial proportion of currently married women in the reproductive age remain largely unmet. The family planning performance is relatively the poorest in these states.

**Table 1.** Use of family planning methods and unmet need for family planning in currently married women aged 15-49 years in selected states and Union Territories of India, 2019-20.

| State/Union Territory | Proportion (%) of currently married women (15-49 years) using a family planning method |  |  |  |  |  |  |  | Unmet need for family planning |  |
| --- | --- | --- | --- | --- | --- | --- | --- | --- | --- | --- |
|  | Any method | Any modern method | Female sterilisation | Male sterilisation | IUD/ PPIUD | Pill | Condom | Injectable | Total | Spacing |
| AN Islands | 65.8 | 57.7 | 39.2 | 0.2 | 3.9 | 3.6 | 9.8 | 0.3 | 13.5 | 6.1 |
| Andhra Pradesh | 71.1 | 70.8 | 69.6 | 0.4 | 0.2 | 0.1 | 0.5 | 0.0 | 4.7 | 2.6 |
| Assam | 60.8 | 45.3 | 9.0 | 0.1 | 2.9 | 27.5 | 4.9 | 0.5 | 11.0 | 4.1 |
| Bihar | 55.8 | 44.4 | 34.8 | 0.1 | 0.8 | 2.0 | 4.0 | 1.1 | 13.6 | 6.1 |
| DN & DD | 68.0 | 59.8 | 41.6 | 0.2 | 2.2 | 3.1 | 11.7 | 0.9 | 11.9 | 5.3 |
| Goa | 67.9 | 60.1 | 29.9 | 0.0 | 2.4 | 2.7 | 23.2 | 0.0 | 8.4 | 4.0 |
| Gujarat | 65.3 | 53.6 | 35.9 | 0.2 | 3.1 | 2.3 | 11.4 | 0.1 | 10.3 | 4.5 |
| Himachal Pradesh | 74.2 | 63.4 | 37.7 | 3.3 | 1.1 | 1.5 | 19.2 | 0.1 | 7.9 | 2.8 |
| Jammu & Kashmir | 59.8 | 52.5 | 21.1 | 0.3 | 5.9 | 9.0 | 11.7 | 3.6 | 7.8 | 3.9 |
| Karnataka | 68.7 | 68.2 | 57.4 | 0.0 | 2.9 | 2.1 | 4.1 | 0.5 | 6.5 | 3.8 |
| Kerala | 60.7 | 52.8 | 46.6 | 0.1 | 1.5 | 0.4 | 3.4 | 0.0 | 12.5 | 7.0 |
| Laddakh | 51.3 | 48.0 | 16.7 | 0.4 | 7.9 | 6.6 | 9.0 | 6.2 | 7.9 | 4.0 |
| Lakshadweep | 52.6 | 30.1 | 20.7 | 0.0 | 1.0 | 1.2 | 4.1 | 0.0 | 12.3 | 8.0 |
| Maharashtra | 66.2 | 63.8 | 49.1 | 0.4 | 1.9 | 1.8 | 10.2 | 0.2 | 9.6 | 3.9 |
| Manipur | 61.3 | 18.2 | 3.7 | 0.0 | 4.9 | 4.4 | 4.8 | 0.1 | 12.2 | 4.7 |
| Meghalaya | 27.4 | 22.5 | 5.6 | 0.0 | 4.4 | 8.3 | 2.7 | 1.1 | 26.9 | 18.3 |
| Mizoram | 31.2 | 30.8 | 13.0 | 0.0 | 2.8 | 12.9 | 1.9 | 0.1 | 18.9 | 12.8 |
| Nagaland | 57.4 | 45.3 | 14.4 | 0.0 | 19.8 | 6.4 | 3.3 | 0.3 | 9.1 | 4.5 |
| Sikkim | 69.1 | 54.9 | 14.5 | 1.7 | 6.2 | 18.2 | 9.3 | 3.5 | 11.9 | 4.9 |
|  | Any method | Any modern method | Female sterilisation | Male sterilisation | IUD/ PPIUD | Pill | Condom | Injectable | Total | Spacing |
| Telangana | 68.1 | 66.7 | 61.9 | 2.0 | 0.5 | 0.8 | 0.8 | 0.1 | 6.4 | 2.8 |
| Tripura | 71.2 | 49.1 | 10.5 | 0.0 | 0.4 | 32.8 | 3.3 | 0.3 | 8.2 | 2.5 |
| West Bengal | 74.4 | 60.7 | 29.4 | 0.1 | 2.2 | 20.3 | 7.0 | 0.7 | 7.0 | 3.0 |

Source: Government of India (2020)
Remarks: AN Islands – Andaman and Nicobar Islands DN & DD – Dadra & Nagar Haveli and Daman and Diu

**Table 2.** Family planning performance indicators in selected states/Union Territories of India, 2019-20.

| State/Union Territory | Performance indexes |  |  |  |  | Rank in performance indexes |  |  |  |  |
| --- | --- | --- | --- | --- | --- | --- | --- | --- | --- | --- |
| | $c_s$ | $c_p$ | $c$ | $q$ | $p$ | $c_s$ | $c_p$ | $c$ | $q$ | $p$ |
| AN Islands | 0.563 | 0.842 | 0.693 | 0.355 | 0.506 | 14 | 12 | 10 | 13 | 11 |
| Andhra Pradesh | 0.216 | 0.971 | 0.503 | 0.020 | 0.151 | 22 | 1 | 19 | 22 | 22 |
| Assam | 0.649 | 0.569 | 0.608 | 0.427 | 0.512 | 9 | 20 | 16 | 10 | 10 |
| Bihar | 0.352 | 0.823 | 0.555 | 0.249 | 0.381 | 18 | 14 | 17 | 18 | 19 |
| DN & DD | 0.571 | 0.864 | 0.708 | 0.331 | 0.496 | 13 | 9 | 8 | 16 | 13 |
| Goa | 0.719 | 0.872 | 0.793 | 0.451 | 0.606 | 3 | 8 | 3 | 9 | 6 |
| Gujarat | 0.519 | 0.862 | 0.676 | 0.355 | 0.499 | 15 | 10 | 12 | 14 | 12 |
| Himachal Pradesh | 0.622 | 0.889 | 0.748 | 0.402 | 0.557 | 10 | 6 | 7 | 11 | 7 |
| Jammu & Kashmir | 0.735 | 0.846 | 0.789 | 0.633 | 0.708 | 2 | 11 | 5 | 5 | 2 |
| Karnataka | 0.715 | 0.955 | 0.829 | 0.183 | 0.428 | 4 | 2 | 1 | 19 | 17 |
| Kerala | 0.290 | 0.895 | 0.537 | 0.135 | 0.291 | 19 | 5 | 18 | 20 | 20 |
| Laddakh | 0.809 | 0.814 | 0.812 | 0.701 | 0.755 | 1 | 15 | 2 | 1 | 1 |
| Lakshadweep | 0.236 | 0.828 | 0.471 | 0.350 | 0.408 | 20 | 13 | 20 | 15 | 18 |
| Maharashtra | 0.694 | 0.897 | 0.791 | 0.254 | 0.473 | 5 | 4 | 4 | 17 | 15 |
| Manipur | 0.233 | 0.330 | 0.279 | 0.660 | 0.442 | 21 | 22 | 22 | 3 | 16 |
| Meghalaya | 0.421 | 0.394 | 0.408 | 0.641 | 0.516 | 16 | 21 | 21 | 4 | 9 |
| Mizoram | 0.574 | 0.681 | 0.626 | 0.490 | 0.555 | 12 | 18 | 15 | 8 | 8 |
| Nagaland | 0.651 | 0.758 | 0.703 | 0.548 | 0.622 | 8 | 16 | 9 | 6 | 4 |
| Sikkim | 0.670 | 0.698 | 0.684 | 0.687 | 0.685 | 6 | 17 | 11 | 2 | 3 |
| Telangana | 0.400 | 0.947 | 0.635 | 0.084 | 0.273 | 17 | 3 | 13 | 21 | 21 |
| Tripura | 0.611 | 0.648 | 0.629 | 0.356 | 0.480 | 11 | 19 | 14 | 12 | 14 |
| West Bengal | 0.651 | 0.881 | 0.760 | 0.495 | 0.618 | 7 | 7 | 6 | 7 | 5 |

**Table 3.** Family planning performance of states/Union Territories from the two-dimensional perspective, 2015-16

| Performance in terms of met demand | Performance in terms of composition of met demand |  |  |  |  |
| --- | --- | --- | --- | --- | --- |
| | Very poor<br>($q < 0.300$) | Poor<br>($0.300 < q \leq 0.550$) | Average<br>($0.550 < q \leq 0.750$) | Good<br>($0.750 < q \leq 0.900$) | Very good<br>($q \geq 0.900$) |
| Very poor<br>($q < 0.300$) | | | Manipur | | |
| Poor<br>($0.550 < c \leq 0.750$) | Andhra Pradesh<br>Kerala | Lakshadweep | Meghalaya | | |
| Average<br>($0.550 < c \leq 0.750$) | Bihar<br>Telangana | AN Islands<br>Assam<br>DA & DD<br>Gujarat<br>Himachal Pradesh<br>Mizoram<br>Nagaland<br>Tripura | Sikkim | | |
| Good<br>($0.750 < c \leq 0.900$) | Karnataka<br>Maharashtra | Goa<br>West Bengal | Jammu & Kashmir<br>Laddakh | | |
| Very good<br>($c \geq 0.900$) | | | | | |

Source: Author's calculations
Remarks: AN Islands – Andaman and Nicobar Islands DA & DD – Dadra & Nagar Haveli and Daman & Diu

**Table 4.** Improvement in family planning performance indexes in selected states/Union Territories of India, 2015-2020.

| State/Union Territory | Improvement between 2015-16 and 2019-20 |  |  |  |  | Improvement rank |  |  |  |  |
| --- | --- | --- | --- | --- | --- | --- | --- | --- | --- | --- |
| | $c_s$ | $c_p$ | $c$ | $q$ | $p$ | $c_s$ | $c_p$ | $c$ | $q$ | $p$ |
| AN Islands | 0.121 | -0.002 | 0.072 | 0.157 | 0.135 | 8 | 16 | 9 | 3 | 6 |
| Andhra Pradesh | 0.054 | -0.006 | 0.048 | 0.000 | 0.010 | 13 | 17 | 14 | 21 | 21 |
| Assam | 0.085 | 0.035 | 0.059 | 0.005 | 0.029 | 9 | 12 | 13 | 19 | 18 |
| Bihar | 0.149 | 0.186 | 0.175 | 0.120 | 0.149 | 7 | 3 | 4 | 7 | 4 |
| DN & DD | 0.217 | 0.092 | 0.171 | 0.137 | 0.159 | 4 | 8 | 5 | 5 | 3 |
| Goa | 0.255 | 0.233 | 0.246 | 0.110 | 0.170 | 2 | 2 | 3 | 8 | 2 |
| Gujarat | 0.047 | 0.096 | 0.069 | 0.106 | 0.097 | 14 | 7 | 10 | 9 | 8 |
| Himachal Pradesh | 0.012 | 0.117 | 0.060 | 0.049 | 0.055 | 16 | 5 | 12 | 10 | 13 |
| Jammu & Kashmir | 0.184 | 0.056 | 0.126 | 0.144 | 0.137 | 5 | 10 | 7 | 4 | 5 |
| Karnataka | 0.430 | 0.038 | 0.288 | 0.122 | 0.207 | 1 | 11 | 1 | 6 | 1 |
| Kerala | 0.007 | 0.000 | 0.005 | 0.032 | 0.030 | 17 | 15 | 19 | 16 | 17 |
| Laddakh | -0.037 | -0.041 | -0.039 | 0.188 | 0.088 | 19 | 21 | 21 | 1 | 9 |
| Lakshadweep | 0.078 | 0.110 | 0.101 | 0.026 | 0.061 | 11 | 6 | 8 | 18 | 11 |
| Maharashtra | 0.055 | -0.008 | 0.027 | 0.039 | 0.040 | 12 | 18 | 15 | 13 | 14 |
| Manipur | -0.054 | 0.175 | 0.064 | 0.037 | 0.059 | 22 | 4 | 11 | 14 | 12 |
| Meghalaya | -0.049 | -0.118 | -0.083 | 0.166 | 0.033 | 20 | 22 | 22 | 2 | 16 |
| Mizoram | -0.013 | -0.015 | -0.014 | 0.033 | 0.012 | 18 | 19 | 20 | 15 | 20 |
| Nagaland | 0.225 | 0.305 | 0.264 | 0.003 | 0.132 | 3 | 1 | 2 | 20 | 7 |
| Sikkim | -0.050 | 0.077 | 0.015 | 0.040 | 0.027 | 21 | 9 | 18 | 12 | 19 |
| Telangana | 0.169 | 0.007 | 0.130 | 0.027 | 0.067 | 6 | 14 | 6 | 17 | 10 |
| Tripura | 0.079 | -0.030 | 0.027 | -0.012 | 0.004 | 10 | 20 | 16 | 22 | 22 |
| West Bengal | 0.031 | 0.013 | 0.023 | 0.041 | 0.034 | 15 | 13 | 17 | 11 | 15 |

In all states/Union Territories, family planning performance has improved between 2015-16 and 2019-20 but the extent of improvement varies widely. The improvement has been the most rapid in Karnataka followed by Bihar but there has been hardly any improvement in performance in Tripura. In 12 other states/Union Territories, the improvement has been less than 20 per cent. The increase in indexes *c* and *q* has also varied widely. The index *c* increased in all but three states/Union Territories. There are, however, only 5 states/Union Territories where the index increased by at least 30 per cent with the increase being the most rapid in Nagaland. In 6 states/Union Territories, there has been only a marginal increase the index. In Meghalaya, Laddakh and Mizoram, however, there has been a decrease in the met demand for family planning. On the other hand, the index *q* increased very rapidly in Karnataka. In Bihar also, the increase in the index has been rapid. The increase in the index *q* implies that the method mix has turned less skewed which indicates that the capacity of the family planning service delivery system in meeting diverse family planning needs of the people has improved. In 9 other states/Union Territories, the index *q* increased substantially but the increase has been marginal in 9 states/Union Territories. By contrast, the index decreased in Tripura and Andhra Pradesh which means that the method mix has turned more skewed. An increase in method skew implies that the capacity of the family planning services delivery system in meeting the diverse family planning needs of the people has decreased.

## Discussions and Conclusions

The evidence available through the latest NFHS 2019-20 suggests that the family planning performance in 22 states/Union Territories of the country remains far from satisfactory in terms of both met demand for modern family planning methods and the structure or the composition of the met demand. It is apparent from the analysis that family planning efforts in the country, either government or non-government, are still not able to meet the diverse family planning needs of the people. The vision 2020 of the Government of India anoints family planning as a critical intervention to reduce maternal and child mortality and morbidity beyond the simple strategy for achieving population stabilization (Government of India, 2014). The present analysis suggests that, to realise this vision, there is a need to substantially reinvigorate family planning efforts, especially, government family planning efforts so that the family planning services delivery system is able to meet the diverse family planning needs of the people. An important requirement, in this direction, is to drastically change the structure of the met demand for modern family planning methods so that there is significant increase in the met demand for modern spacing methods. Some states and Union Territories of the country appear to have been able to achieve such an increase. However, in most of the states/Union Territories, the structure of the met demand of family planning remains heavily skewed towards permanent methods so that the met demand for modern spacing methods remains unacceptably low. At the policy level, the emphasis on shifting the focus of family planning efforts from birth limitation to birth planning have been oft-repeated but the latest available evidence suggests that progress in this direction remains lethargic in most of the states/Union Territories.

Although, policies and programmes to meet the diverse family planning needs of the people in India are formulated at the national level, yet the responsibility of implementing these policies and programmes essentially, rests with constituent states and Union Territories of the country. The variation in both met demand for modern family planning method and the structure of the met demand across states/Union Territories suggests that family planning performance is conditioned by factors that are specific to the state/Union Territory. At present, very little is known about these factors but an understanding of these factors is critical to improving the family planning performance in the country in the context of meeting the family planning needs of the people.

More and more states and Union Territories in India are now achieving the replacement fertility. It is now the time that family planning is now pursued as a development strategy rather than a health intervention. Potential benefits of family planning include economic development, improvement in maternal and child health, educational advancement, empowerment of women, and environmental protection (Bongaarts et al, 2012; Cleland et al, 2006). It is a proven, cost-effective intervention for preventing mother-to-child transmission of HIV (Reynolds et al, 2005; Reynolds et al, 2006; Reynolds et al, 2008; Stover et al, 2003) and can protect against both unintended pregnancy and sexual transmission of HIV (Wilcher et al, 2009). Benefits of family planning impact all the 17 Sustainable Development Goals (Starbird et al, 2016). It is estimated that ‘every dollar invested in family planning saves four dollars in other health and development areas” (Toure et al, 2012; Frost et al, 2008). Reinvigorating family planning efforts is, therefore, the need of the time for India in its quest for rapid social and economic development.

## Data Availability

All data used in the analysis are in public domain.

